# Triaging clients at risk of disengagement from HIV care: Application of a predictive model to clinical trial data in South Africa

**DOI:** 10.1101/2024.08.05.24311488

**Authors:** Mhairi Maskew, Shantelle Smith, Lucien De Voux, Kieran Sharpey-Schafer, Thomas Crompton, Ashley Govender, Pedro Pisa, Sydney Rosen

## Abstract

**Background:** To reach South Africa’s targets for HIV treatment and viral suppression, retention on antiretroviral therapy (ART) must increase. Much effort and resources have been invested in tracing those already disengaged and returning them to care programs with mixed success. Here we aim to successfully identify ART clients at risk of loss from care prior to disengagement.

**Methods and Findings:** We applied a previously developed machine learning and predictive modelling algorithm (PREDICT) to routinely collected ART client data from the SLATE I and SLATE II trials, which evaluated same-day ART initiation in 2017-18. Using a primary outcome of an interruption in treatment (IIT), defined as missing the next scheduled clinic visit by >28 days, we investigated the reproducibility of PREDICT in SLATE datasets. We also tested two risk triaging approaches: 1) threshold approach classifying individuals into low, moderate, or high risk of IIT; and 2) archetype approach identifying subgroups with characteristics associated with risk of ITT. We report associations between risk category groups and subsequent IIT at the next scheduled visit using crude risk differences and relative risks with 95% confidence intervals. SLATE datasets included 7,199 client visits for 1,193 clients over ≤14 months of follow-up. The algorithm achieved 63% accuracy, 89% negative predictive value, and an area under the curve of 0.61 for attendance at next scheduled visit, similar to previous results using only medical record data. The threshold approach consistently and accurately assigned levels of IIT risk for multiple stages of the care cascade. The archetype approach identified several subgroups at increased risk of IIT, including those late to previous appointments, those returning after a period of disengagement, those living alone or without a treatment supporter. Behavioural elements of the archetypes tended to drive risk of treatment interruption more consistently than demographics; e.g. adolescent boys/young men who attended visits on time experienced lowest rates of treatment interruption (10%, PREDICT datasets and 7% SLATE datasets), while adolescent boys/young men returning after previously disengaging from care had highest rates of subsequent treatment interruption (31%, PREDICT datasets and 40% SLATE datasets).

**Conclusion:** Routinely collected medical record data can be combined with basic demographic and socioeconomic data to assess individual risk of future treatment disengagement using machine learning and predictive modelling. This approach offers an opportunity to intervene prior to and potentially prevent disengagement from HIV care, rather than responding only after it has occurred.

## INTRODUCTION

With the successful expansion of universal access to HIV treatment around the world, retaining persons living with HIV in lifelong antiretroviral therapy (ART) has emerged as one of the most important challenges to HIV epidemic control [1]. For those who disengage from care (i.e. are not retained), the most common intervention continues to be after-the-fact tracking and tracing efforts, in which healthcare workers attempt to contact disengaged ART clients and encourage and/or assist them to return to care. These efforts have had mixed results, in terms of achieving re-engagement in care [2–8].

A major drawback to all tracking and tracing programs is that they can only intervene after a person disengages from care. Little is done to distinguish those at higher risk of dropping out of care in advance, before disengagement occurs. Instead, the same advance support is offered to all, regardless of risk level. A strategy for identifying individuals at high risk of disengagement before they interrupt care would allow interventions to be targeted to those in need up front, before any damage is done, while conserving the resources that might otherwise be expended on low risk clients who require little or no intervention to remain in care [9]. To put such a strategy into practice, both accurate pre-interruption risk triaging and a practical, low-cost tool that frontline healthcare workers can use to identify ART clients for differing levels of retention support and interventions are needed.

A number of previous efforts have been made to predict risks of poor outcomes among people living with HIV [10–16]. While several models include basic demographic characteristics such as age and sex and clinical history such as baseline CD4 count to predict risk, the mechanisms driving risk within demographic subgroups at higher risk of disengagement than their age/sex peers remain unclear. Other characteristics that predict risk may be important to identify because within virtually any “risky” age/sex stratum, such as young men [17,18], a majority of individuals remain low risk and achieve good outcomes without intervention. In at 2018 population survey in KwaZulu Natal South Africa, for example, young men aged 15-29 were the highest risk age/sex group identified, but more than half of them (51.5%) were virally suppressed[19].

We previously applied machine learning and predictive algorithms [20] to routinely collected longitudinal HIV phenotypic and clinical outcome data from the South African HIV treatment programme, one of the largest globally [21]. The PREDICT model aimed to identify those at risk of a near-term interruption in treatment (IIT), defined as missing their next scheduled clinic visit by more than 28 days. To move beyond the age/sex and visit history characteristics that are currently routinely collected in electronic medical records, we reproduced this model in a smaller South African HIV clinical trial dataset from the SLATE trials [15,22] containing socioeconomic indicators. We then utilized the output for two risk score triaging approaches to identify those at risk for disengagement from care: 1) a threshold approach to segment populations into risk groups; and 2) a series of archetypes characterizing social and behavioral client profiles. Here, we describe the development of these approaches and estimate associations with risk of disengagement from care, providing the basis for future development of a practical, point-of-care risk triaging tool.

## METHODS

### Population and data sources

The two approaches to risk triaging were developed using output derived from two machine learning models. The first, PREDICT (Prioritizing Retention Efforts using Data Intelligence and Cohort Targeting) [23], was initially trained and tested on routinely collected, anonymized, longitudinal medical record data from clients accessing HIV care and treatment at public sector treatment sites in Mpumalanga and the Free State between January 2016 and December 2018. These records contain information on clients’ clinical and antiretroviral treatment histories, including scheduled and attended clinical visits and laboratory test results, and basic demographic characteristics (age and sex). On average, PREDICT correctly identified two out of three clients who missed their next scheduled clinic visit. The model was recently validated in a different population and geographic setting in South Africa and demonstrated almost identical performance metrics [24].

For the second model, the SLATE model, we used client survey and medical record data collected for the SLATE I and SLATE II trials, which were randomized evaluations of a clinical algorithm to determine eligibility for same-day initiation of ART at three primary healthcare facilities in Gauteng Province [15,22,25]. SLATE enrolled non-pregnant adults who presented at the study clinics for any kind of HIV care, including diagnosis, and were not yet on ART. Participants completed a baseline survey that included demographic and socioeconomic characteristics, HIV testing and treatment history, and social indicators including disclosure status. Participants were then passively followed up for 14 months after study enrolment through clinic medical records observing scheduled and attended clinic visits at the study sites.

### Study outcomes

The primary outcome of interest was retention in HIV care. We considered a client to be retained in care if a clinic visit was observed before or within 28 days of the next scheduled appointment date in that client’s medical record[26]. Conversely, we defined a client to have experienced an interruption in treatment (IIT) when a client did not attend a clinic visit within 28 days of their scheduled appointment. We restricted the analysis to visits scheduled a minimum of three months prior to the database censor date to allow for one month to meet the outcome definition and a further two months to allow for capturing visit data into the EMR. All raw data available in the source datasets were considered as potential predictors of IIT. These included data characterizing client demographics, HIV testing and ART treatment history, socio-economic indicators (employment, income), disclosure, drug regimen data, visit history and patterns, and ART monitoring laboratory test results.

### Model building and performance

Both the PREDICT and SLATE models used the AdaBoost (adaptive boosting binary classification) algorithm from scikit-learn [27]. The model building and validation process is detailed in Supplementary File 1 and also described elsewhere [20,24]. In short, each of the source datasets is split into training and test sets. Training sets are datasets with known exposure and outcome variables used in machine learning approaches to allow the algorithm to “learn” the predictive importance of exposure variables in terms of correctly classifying each specified outcome. For test sets, the exposure variables are separated from the outcome variables (unseen) and given to the final classifier algorithm. The model is tested on this unseen data set by generating predicted outcomes for each observed visit using the predictor variables from the unseen test set. In this way, the model produces an overall predicted risk score for each visit that indicates the likelihood that the next scheduled visit will not be attended on time and will be classified as an interruption in treatment. These predicted outcomes are then compared to the known outcomes in the test set and the model is scored according to standard test performance metrics (sensitivity, specificity, positive and negative predictive value and area under the curve). To determine the value of the additional variables added to the medical record data using the SLATE baseline survey questionnaires, we estimated model performance metrics when restricting the SLATE model to the set of variables that were available to the PREDICT model (i.e. data from routinely collected medical records only).

### Risk score triaging

We next adopted two approaches to create a risk score triaging system to identify groups at risk of IIT. Output from both the SLATE and PREDICT models were used in each of the two risk-score triaging systems and are presented stratified by source model for comparison throughout, with the exception of situations where the model did not contain variables required to classify risk groups or profiles. The two approaches are described below and compared in Panel 1.

**Panel 1:** Comparison of threshold and archetype approaches to risk triaging.

| PROPERTY | THRESHOLD APPROACH | ARCHETYPE APPROACH |
| --- | --- | --- |
| General approach | Classifies clients into groups at low (green), moderate (yellow) or high (red) risk of outcome based on predictive model scores | Identifies sub-clusters of client populations with characteristics associated with risk of outcome |
| Automation | Can be automated into predictive model output; integrated into EMR | Currently involves manual application of clustering characteristics identified by machine learning model. Automation through EMR integration may be possible in future versions |
| Applicability | Applicable to any dataset and population | Specific to population context and characteristics |
| Intuitiveness or understandability | Requires score to be calculated beforehand and may not always be consistent with clinician assessment of risk | Intuitive, easy to apply with clients at point of care |
| Identification of drivers of risk | Mechanism behind risk not readily apparent | Some underlying driver of risk can be ascertained through group characteristics |
| Intervention mapping | Less directly useful for intervention mapping | May offer opportunities for direct intervention mapping |

The *threshold approach* grouped the final predicted risk scores assigned to each visit by the PREDICT or SLATE model into three pre-set categories: visits with the lowest 50% of scores were assigned a “green” or low risk category; visits with the middle 40% of scores were assigned a “yellow” or moderate risk category; and visits scoring in the highest 10% of risk scores were assigned a “red” or high-risk category. We then considered the visit observed immediately after a “scored” visit (hereafter called “next visit”) and classified this as IIT or not, based on whether the next visit occurred within 28 days of its scheduled date. The proportion of next visits classified as IIT was then estimated for each risk triage category.

The *archetype approach* used characteristics identified by each model as important predictors of missed visits. The PREDICT model considered demographic and visit history characteristics available in the routine EMR datasets while the SLATE model used social, economic, and HIV treatment experience features collected as part of the clinical trial enrolment survey, in addition to demographic and visit history characteristics from the EMR. ART clients were then grouped together into subgroups with a shared set of characteristics, creating distinct sub-population profiles or archetypes.

Using feature importance tables from the predictive modeling, features were next paired into different configurations. (For example, combining responses to the questions ‘Has the client disclosed their status?’ and ‘Does the client have enough information to start ART?’ yields four configurations: not disclosed and not enough information; have disclosed and not enough information; not disclosed and have enough information; have disclosed and have enough information.) These archetypes were then used to isolate the subgroups of the population where those two or three features were key in determining their risk of an interruption in treatment. Any logically invalid or very small subgroups were removed. Finally, the rate of IIT was calculated within each sub-group (or configuration) and all subgroups’ IIT rates were then compared to the whole population’s baseline IIT rate. The groupings that had the largest positive or negative differences from baseline were identified as potential archetypes of interest.

### Statistical analysis

For the threshold approach, we first used simple frequencies and proportions to describe the overall number and distribution of visits triaged into each risk category (green, yellow and red groups). We stratified these descriptive statistics by age, gender, and time on ART. Next, we estimated the crude relative risk (RR) and corresponding 95% confidence interval (CI) for IIT at next visit stratified by current visit risk triage category, with the green “low risk” group as reference. The analytic approach to the client archetypes was similar. We first described the overall frequency and distribution of visits by clients characterized into each archetype, stratified into age and gender clusters and by time on ART. We then estimated the crude relative risk and corresponding 95% confidence interval of missing a scheduled visit for comparing each archetype to the archetype with the lowest perceived risk of IIT.

### Ethics statement

All analyses of de-identified data from human subjects were approved by and carried out in accordance with relevant guidelines and regulations as set out by the Human Research Ethics Committee of the University of the Witwatersrand (Medical). This study involved secondary analysis of two data sources: 1) deidentified data collected as part of routine care, for which the requirement for individual patient consent was waived by the Human Research Ethics Committee of the University of the Witwatersrand for protocols M140201 and M210472 during the study approval; and 2) de-identified clinical trial collected as part of the SLATE I and SLATE II trials (Clinicaltrials.gov registration NCT02891135). Both studies were approved by the Human Research Ethics Committee of the University of the Witwatersrand (Medical) and the institutional review board of Boston University Medical Campus. All SLATE study participants provided written informed consent.

## RESULTS

### Characteristics of study participants and model performance metrics

The two source data sets are described in Table 1. The original PREDICT model data set utilized routinely collected, anonymized, longitudinal data from >460,000 clients accessing HIV care and treatment during >4.6M visits at public sector treatment sites in Mpumalanga and the Free State between January 2016 and December 2018. The SLATE trials provided a total of 1,193 patient records containing 7,199 clinic visits in Gauteng. Participant characteristics are summarized in Table 1. We note that the SLATE study population differed substantially from the original PREDICT model dataset by the distribution of stage of HIV care journey. The original PREDICT data set included visits across all stages of care with a median duration on ART of approximately 5 years. SLATE study participants, in contrast, were all enrolled at ART initiation and followed up for a maximum duration of 14 months. Pregnant women were also excluded from the SLATE studies but included in the PREDICT datasets.

**Table 1:** Characteristics of the study population.

| Characteristic | PREDICT model | SLATE model |
| --- | --- | --- |
| Data source | Routinely collected EMR data | Clinical trial data supplemented with EMR data |
| Setting | Ehlanzeni District (Mpumalanga) and Thabo Mofutsanyana District (Free State) | City of Johannesburg and Ekurhuleni Districts (Gauteng) |
| Facility profile (%) |  |  |
| Urban | 52% | 67% |
| Peri-urban | 9% | 33% |
| Rural | 37% | 0% |
| Missing | 2% | 0% |
| Client sample size | 463,418 clients | 1,156 clients |
| Visit sample size | 4,663,816 visits | 7,199 visits |
| Current age (median, IQR) | 39 years (27-49) | SLATE: 35 years (29-41) |
| % Female | 68% | 64% |
| Prevalent pregnancy | 2% | 0% (pregnant women were excluded) |
| Time on ART at entry to cohort (median, IQR) | 62 months (30-93 months) | 0 months (all newly initiating or re-initiating clients) |
| Maximum follow up duration | 36 months | 14 months |
| Variables with most predictive power in the final model | % Visits attended >3 days late<br># Times >28 days late<br># Visits at this facility<br># VL tests done<br>Months since first visit<br>Months since last visit<br>Current age<br>Day of month of next appointment<br>Viral load value (copies/mm<sup>3</sup>)<br>Day of week of next appointment<br>Visits on regimen<br>Sex (M/F)<br># Missed months | Time on ART<br>Appointment month<br>Time since last viral load test<br>Viral load value (copies/mm<sup>3</sup>)<br>% Visits attended >3 days late<br>CD4 count at screening<br>Current age<br>Travel time to clinic<br>Total # TB symptoms<br>Year first tested positive<br># Other people living with the client |

The SLATE data set was divided into a training set of 5,759 visits by 872 clients and a test set of 1,440 visits by 668 clients. In total, 13.5% of visits in the training set and 14.0% visits in the test set were observed to occur >28 days after the scheduled visit date. The algorithm investigated 239 exposure variables in total, including the additional demographic and socioeconomic variables from the SLATE baseline questionnaires. The full set of exposure variables was then reduced to a parsimonious model containing the top 11 exposure features with the most predictive power: time on ART, appointment month, time since last viral load (VL) test, VL test result, proportion of visits attended >3 days late, CD4 count at screening, age, travel time to clinic, total number of TB symptoms, year first tested positive, and number of others living with client in their house.

The SLATE model achieved an accuracy of 63%, specificity of 64%, and negative predictive value of 89% (Table 2), comparable to the original (and much larger) PREDICT dataset which achieved an accuracy of 66%, specificity of 67%, and negative predictive value of 94%.

**Table 2:**
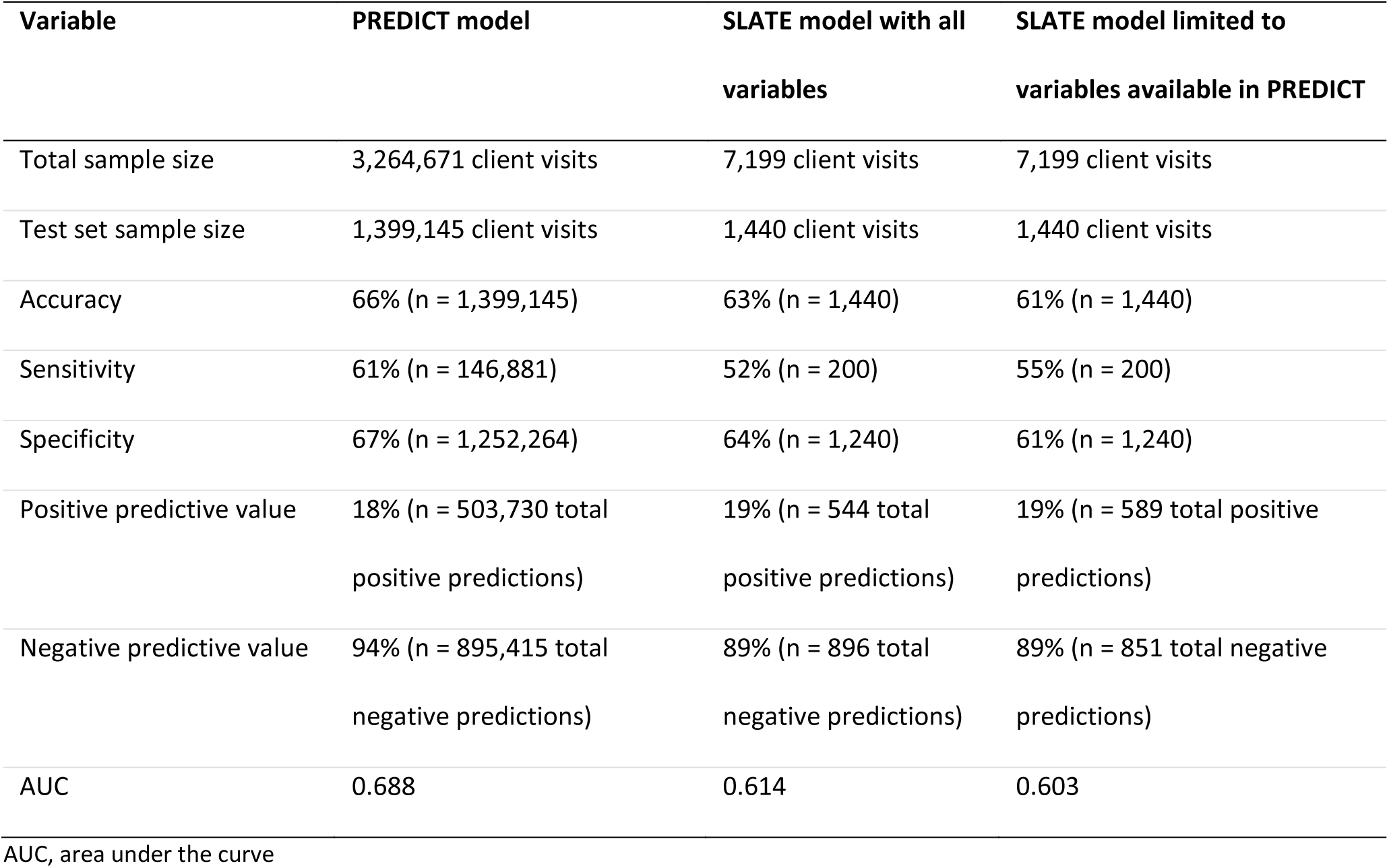
Comparison of SLATE model performance metrics to original PREDICT model.

| Variable | PREDICT model | SLATE model with all variables | SLATE model limited to variables available in PREDICT |
| --- | --- | --- | --- |
| Total sample size | 3,264,671 client visits | 7,199 client visits | 7,199 client visits |
| Test set sample size | 1,399,145 client visits | 1,440 client visits | 1,440 client visits |
| Accuracy | 66% (n = 1,399,145) | 63% (n = 1,440) | 61% (n = 1,440) |
| Sensitivity | 61% (n = 146,881) | 52% (n = 200) | 55% (n = 200) |
| Specificity | 67% (n = 1,252,264) | 64% (n = 1,240) | 61% (n = 1,240) |
| Positive predictive value | 18% (n = 503,730 total positive predictions) | 19% (n = 544 total positive predictions) | 19% (n = 589 total positive predictions) |
| Negative predictive value | 94% (n = 895,415 total negative predictions) | 89% (n = 896 total negative predictions) | 89% (n = 851 total negative predictions) |
| AUC | 0.688 | 0.614 | 0.603 |

AUC, area under the curve

When restricting the SLATE model to the set of variables that were available to the PREDICT model (i.e. data from routinely collected medical records only), the performance of SLATE model demonstrated little change from results obtained using all variables available in the SLATE datasets: 61% accuracy, 61% specificity, and 89% negative predictive value. Results using an alternate model building approach (gradient boosting) to the SLATE data are provided in Supplementary Table 1 for comparison. Hereafter, all results from the SLATE datasets refer to the full model using all variables available in the SLATE datasets unless otherwise stated.

### Results for threshold approach to risk triaging

As explained above, for the threshold approach, results from the predictive models were used to assign a final predictive risk score to every observed visit in the PREDICT and SLATE datasets. These scores were then grouped into centile brackets: the visits with the lowest 50% of scores were assigned a “green” or low risk category; the middle 40% were assigned a “yellow” or moderate risk category; and visits with the highest 10% of scores were assigned a “red” or high-risk category. We then considered the visit observed immediately after a “scored” visit (hereafter called “next visit”) and classified these as IIT or not based on whether the next visit occurred within 28 days of its scheduled date.

In total, 11% of all visits observed in the PREDICT datasets were classified as IIT (n=146,881 visits). The IIT rate observed for visits in the SLATE datasets was slightly higher, at 14% (n=200 visits; Table 3). Rates of IIT at next visit increased in a linear fashion with the increasing predicted risk threshold categories for current visit. Compared to green “low risk” visits in the PREDICT datasets, visits classified in a yellow “moderate risk” group were twice as likely to be followed by a treatment interruption (13% IIT at next visit in yellow group versus 6% IIT at next visit for green group; RR=2.17; 95% CI 2.14-2.19), while the red “high risk” triage visits were more than 4 times as likely to be followed by a treatment interruption at next visit compared to visits classified as green (26% IIT at next visit in red group versus 6% IIT at next visit in green group; RR = 4.33; 95% CI 4.28-4.39). Results were similar using the SLATE datasets.

**Table 3:** Proportion visits with IIT at next scheduled visit stratified by current visit risk triaging classification and time on ART (threshold approach)

| Risk triaging classification at current visit | ALL VISITS |  | FIRST VISIT AFTER INITIATION<br>(n=41,751) |  | 0-6 MONTHS ON ART<br>(n=194,469) |  | 7-12 MONTHS ON ART<br>(n=129,764) |  |
| --- | --- | --- | --- | --- | --- | --- | --- | --- |
|  | IIT at next visit | Relative risk (95% CI) | IIT at next visit | Relative risk (95% CI) | IIT at next visit | Relative risk (95% CI) | IIT at next visit | Relative risk (95% CI) |
| *PREDICT model datasets* |  |  |  |  |  |  |  |  |
| **ALL**<br>(n=1,399,145) | **11%**<br>(n=146,881) |  | **19%**<br>(7,979/41,751) |  | **11%**<br>(19,583/194,469) |  | **10%**<br>(12,970/129,764) |  |
| **GREEN**<br>(n=699,573) | **6%**<br>(n=41,974) | Ref | **13%**<br>(2,714/20,878) | Ref | **6%**<br>(5,835/97,245) | Ref | **5%**<br>(3,244/64,889) | Ref |
| **YELLOW**<br>(n= 559,658) | **13%**<br>(n=72,756) | RR=2.17<br>(2.14-2.19) | **21%**<br>(3,068/14,609) | RR=1.62<br>(1.54-1.69) | **12%**<br>(8,165/68,044) | RR=2.00<br>(1.94-2.07) | **11%**<br>(4,994/45,404) | RR=2.20<br>(2.11-2.30) |
| **RED**<br>(n = 139,915) | **26%**<br>(n=36,378) | RR=4.33<br>(4.28-4.39) | **33%**<br>(2,067/6,264) | RR=2.54<br>(2.42-2.67) | **21%**<br>(6,128/29,180) | RR=3.50<br>(3.38-3.62) | **23%**<br>(4,478/19,471) | RR=4.60<br>(4.41-4.80) |
| *SLATE model datasets* |  |  |  |  |  |  |  |  |
| **ALL**<br>(n=1,440) | **14%**<br>(n=200) |  | **19%**<br>(34/155) |  | **15%**<br>(114/791) |  | N/A |  |
| **GREEN**<br>(n=720) | **10%**<br>(n=75) | Ref | **18%**<br>(14/78) | Ref | **10%**<br>(41/396) | Ref |  |  |
| **YELLOW**<br>(n= 576) | **14%**<br>(n=81) | RR=1.41<br>(1.04-1.89) | **24%**<br>(15/62) | RR=1.35<br>(0.71-2.58) | **18%**<br>(56/316) | RR=1.79<br>(1.23-2.60) |  |  |
| **RED**<br>(n = 144) | **31%**<br>(n=44) | RR=3.13<br>(2.25-4.33) | **33%**<br>(5/15) | RR=1.86<br>(0.79-4.38) | **22%**<br>(17/79) | RR=2.13<br>(1.27-3.56) |  |  |

Ref, Reference population

The rate of IIT at next scheduled visit also differed by time on ART for both the PREDICT and SLATE datasets (Figure 1 and Table 3). Risk of IIT at the next scheduled visit after ART initiation was nearly double that of the periods 0-6 months or 7-12 months on ART (19% versus 10%, respectively). Visits that occurred during month 7-12 after ART initiation and were classified as green had the lowest rates of IIT at next scheduled visit (5%) while first visits after initiation that were classified as red were followed by the highest rates of treatment interruption at next scheduled visit (33%). Within the first 6 months on treatment, visits classified as red were more than three times as likely to be followed by a treatment interruption than visits classified as green (RR 3.50; 95% CI 3.38-3.62); during months 7-12 red visits were more than four times as likely to be followed by an IIT at next scheduled visit as were green visits in the same period (5% vs. 23%; RR=4.60; 95% CI 4.41-4.80). Models generally performed somewhat better in terms of accuracy, sensitivity, specificity and AUC for the full period 0-6 months on ART compared to predictions made only for the first visit (Supplementary Table 2).

**Figure 1:**
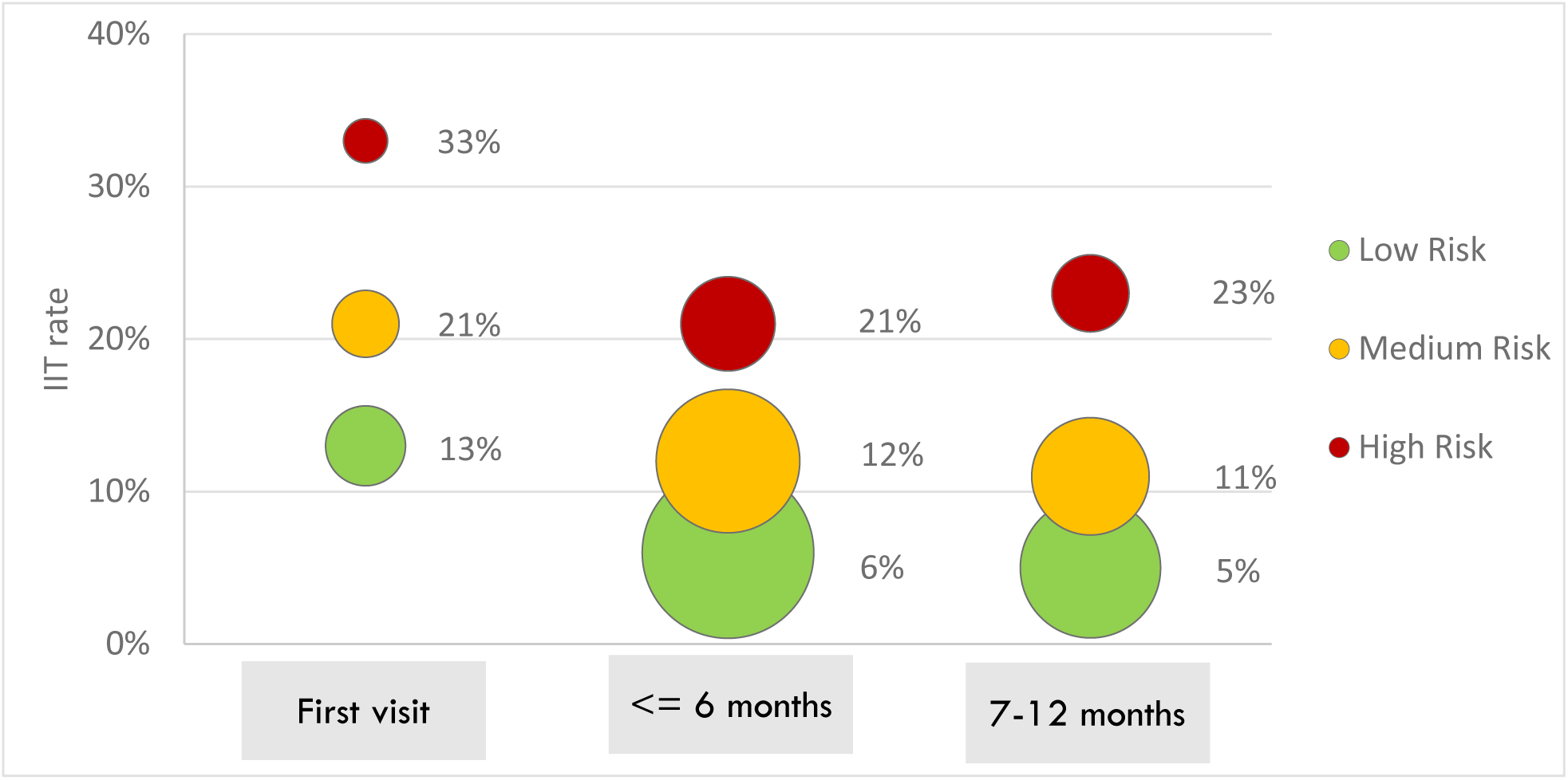
Proportion of visits classified as IIT stratified by risk threshold approach and time on ART (PREDICT model data)

### Results for the archetype approach to triaging

Using characteristics identified by the SLATE model as important predictors of missed visits, we defined archetype profiles across three categories: 1) demographic archetypes based on age and gender; 2) behavioral archetypes based on visit attendance; and 3) social-behavioral archetypes based on client characteristics. In Panel 2, we define archetypes within each category, giving each archetype a descriptive label. As the PREDICT datasets did not contain several of the variables needed to define the socio-behavioural archetypes, these are reported for the SLATE datasets only. The SLATE datasets did not observe movement across facilities and so the behavioural archetype “Shopper” is reported for PREDICT only. All other archetypes are reported for both datasets.

**Panel 2:** Archetype approach definitions.

| Category and archetype | Description | Dataset source | Data collection period |
| --- | --- | --- | --- |
| **Demographic archetypes** |  |  |  |
| Adult female | Female client aged >25 years at date of visit | SLATE and PREDICT | At ART initiation and each follow up visit |
| Adult male | Male client aged >25 years at date of visit | SLATE and PREDICT | At ART initiation and each follow up visit |
| Adolescent girls and young women | Female client aged between 15 and 25 years at date of visit | SLATE and PREDICT | At ART initiation and each follow up visit |
| Adolescent boys and young men | Male client aged between 15 and 25 years at date of visit | SLATE and PREDICT | At ART initiation and each follow up visit |
| **Behavioral archetypes** |  |  |  |
| Prompt and loyal | Visit attended on time and only attended care at this facility | SLATE and PREDICT | At each follow up visit |
| Late twice | The previous two visits were attended after the scheduled appointment date | SLATE and PREDICT | At each follow up visit |
| Shopper, no number | Has attended at other facilities and no phone number on file | PREDICT | At each follow up visit |
| Returning after disengagement | At least one prior visit was attended >28 days late | SLATE and PREDICT | At re-initiation visit |
| ***Socio-behavioral archetypes*** |  |  |  |
| Super green | Punctual visit attendance, doesn't live alone | SLATE | At ART initiation and each follow up visit |
| Employed youth at payday | Age 18-29, identify as employed AND next visit scheduled <7 days from payday | SLATE | At ART initiation and each follow up visit |
| Prior test and prompt | Has a history of HIV testing (before testing positive) AND regularly prompt for visits | SLATE | At ART initiation and each follow up visit |
| Lone ranger | Lives alone or with 1 other person AND lives more than 20 mins away | SLATE | At ART initiation |
| Unexpected and unsupported | Was not planning to test for HIV today AND lives alone/with 1 other person | SLATE | At ART initiation |
| Disillusioned disclosers | Identifies as having HIV info, has disclosed, lives alone or with 1 other person | SLATE | At ART initiation |
| Live close, always late | Lives <20 mins from clinic but is also regularly late for appointments | SLATE | At ART initiation and each follow up visit |
| Prepared and late | Prepared to start ART today, has tested before but is late to appointments | SLATE | At ART initiation and each follow up visit |

Adult females comprised the largest demographic group in both the SLATE and PREDICT datasets (57%) and between 10% and 13% of visits made by adult females were classified as IIT (Table 4). Adult males made up nearly a third of clients in both datasets, with 12-14% of visits made by adult males classified as IIT. Few adolescent or young men and women (8%) were observed in the SLATE datasets, as the trials enrolled participants >18 years of age only. Despite being one of the smallest population groups, adolescent girls and young women (AGYW) demonstrated the highest rates of IIT across the demographic archetypes (15% in SLATE and 16% in PREDICT) and were more likely to have a treatment interruption compared to adult women in both the PREDICT (RR=1.52; 95% CI 1.49-1.55) and SLATE datasets (1.15; 95% CI 0.91-1.46).

**Table 4:** Proportion of visits classified as IIT stratified by archetype triaging approach.

| ARCHETYPES | SLATE DATA (N=7,199 VISITS) |  |  | PREDICT DATA (N=925,639 VISITS)** |  |  |
| --- | --- | --- | --- | --- | --- | --- |
|  | Visits | IIT (n, %) | RR (95% CI)* | Visits | IIT (n, %) | RR (95% CI)* |
| **Demographic archetypes (variables available in both SLATE and PREDICT models)** |  |  |  |  |  |  |
| Adult females | 4,141 (57%) | 555 (13%) | Ref | 572,154 (57%) | 58,271 (10%) | Ref |
| AGYW*** | 434 (6%) | 67 (15%) | 1.15 (0.91-1.46) | 70,045 (7%) | 10,864 (16%) | 1.52 (1.49-1.55) |
| ABYM*** | 146 (2%) | 19 (13%) | 0.97 (0.63-1.49) | 10,444 (10%) | 1,304 (13%) | 1.23 (1.17-1.29) |
| Adult males | 2,478 (34%) | 893 (14%) | 1.02 (0.9 -1.14) | 272,996 (27%) | 31,279 (12%) | 1.20 (1.19 -.21) |
| **Behavioral archetypes (variables available in both SLATE and PREDICT models)** |  |  |  |  |  |  |
| Prompt and loyal | 1,552 (22%) | 227 (15%) | Ref | 652,595 (65%) | 59,099 (9%) | Ref |
| Late twice | 854 (12%) | 155 (18%) | 1.24 (1.03-1.50) | 97,986 (10%) | 15,932 (16%) | 1.80 (1.76-1.83) |
| Shopper no-number |  | N/A |  | 68,087 (7%) | 11,360 (17%) | 1.91 (1.87 -1.94) |
| Returning after disengagement | 861 (12%) | 169 (20%) | 1.34 (1.12-1.61) | 37,404 (4%) | 9,280 (25%) | 2.74 (2.69 -2.80) |
| **Socio-behavioral archetypes (variables available in SLATE model only)** |  |  |  |  |  |  |
| Super green | 2,313 (32%) | 239 (10%) | Ref |  |  |  |
| Employed youth at payday | 301 (4%) | 35 (12%) | 1.13 (0.81-1.57) |  |  |  |
| Prior test and prompt | 1,789 (25%) | 228 (13%) | 1.23 (1.04-1.46) |  |  |  |
| Lone ranger | 1,478 (21%) | 221 (15%) | 1.45 (1.22-1.72) |  |  |  |
| Unexpected and unsupported | 817 (11%) | 120 (15%) | 1.42 (1.16-1.74) |  |  |  |
| Disillusioned disclosers | 1,194 (17%) | 184 (15%) | 1.49 (1.25-1.78) |  |  |  |
| Live close but always late | 986 (14%) | 167 (17%) | 1.64 (1.36-1.97) |  |  |  |
| Prepared and late | 501 (7%) | 93 (19%) | 1.80 (1.44-2.24) |  |  |  |

\* RR = Relative risk, reported with 95% confidence interval
\*\* Data restricted to visits within the first 6 months on ART
\*\*\* AGYW = adolescent girls and young women, ABYM = adolescent boys and young men

Several of the identified behavioral archetypes were also at increased risk of IIT at next visit compared to their reference groups. Clients who had been late for at least two prior visits were more likely to have an IIT at next visit compared to all adult women (Table 4) in both datasets. Those who were returning after previously disengaging from care were at the highest risk of IIT compared to adult females (RR = 2.44; 95% CI 2.40-2.48 in PREDICT and RR=1.46; 95% CI 1.25-1.71 in SLATE). When combining social and behavioural characteristics (SLATE data only), the client archetypes least likely to have an IIT at next visit were those who attended prior visits on time, were young and employed, and had a history of previous HIV testing. Those who lived alone, did not have a treatment supporter, or were not expecting to start HIV treatment at initiation were at increased risk of having a treatment interruption. Compared to those who attend visits on time and don’t live alone, youth who reported being employed and had a visit scheduled within 7 days of payday were at a somewhat increased risk of a subsequent treatment interruption (12% IIT; RR = 1.13; 95% CI 0.81-1.57)

We also stratified the behavioural and socio-behavioural archetypes by age and gender and noted varying risk for different substrata of the population (Table 5). In particular, we noted that the behavioural elements (visit attendance) of the archetypes tended to drive the risk of treatment interruption more consistently than the basic demographic elements. For example, adolescent boys and young men who attended visits on time experienced one of the lowest rates of treatment interruption (10%, PREDICT datasets and 7% SLATE datasets), while adolescent boys and young men who had returned after previously disengaging in care were the group with highest rates of subsequent treatment interruption (31%, PREDICT datasets and 40% SLATE datasets). Similarly, adolescent girls and young women returning after a period of disengagement were 3.5 times more likely to have a treatment interruption when compared to adult females (RR=3.50; 95% CI 3.32-3.68; PREDICT datasets). In fact, even a visit history of attending late twice among adolescent girls and young women was associated with a subsequent treatment interruption (21% IIT, RR=2.50 (95% CI 2.39-2.61) in PREDICT data and 26% IIT, RR =1.93 (95% CI 1.23-3.05) in SLATE datasets) compared to all adult females. Other socio-behavioural archetypes associated with increases in risk for subsequent treatment interruption regardless of demographic profile included archetypes characterized by limited or no social support at home and living alone and/or at a far distance from the clinic.

**Table 5:**
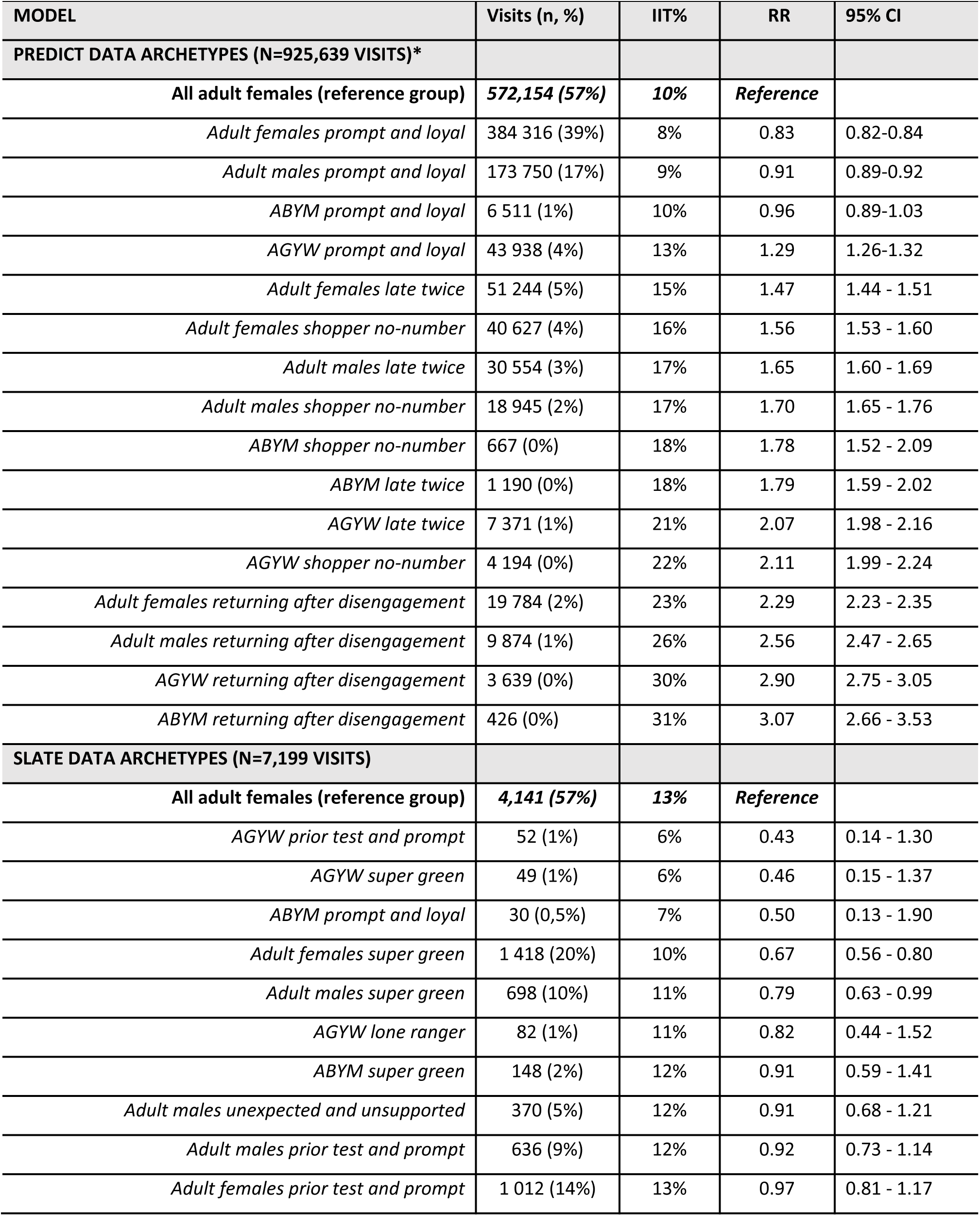
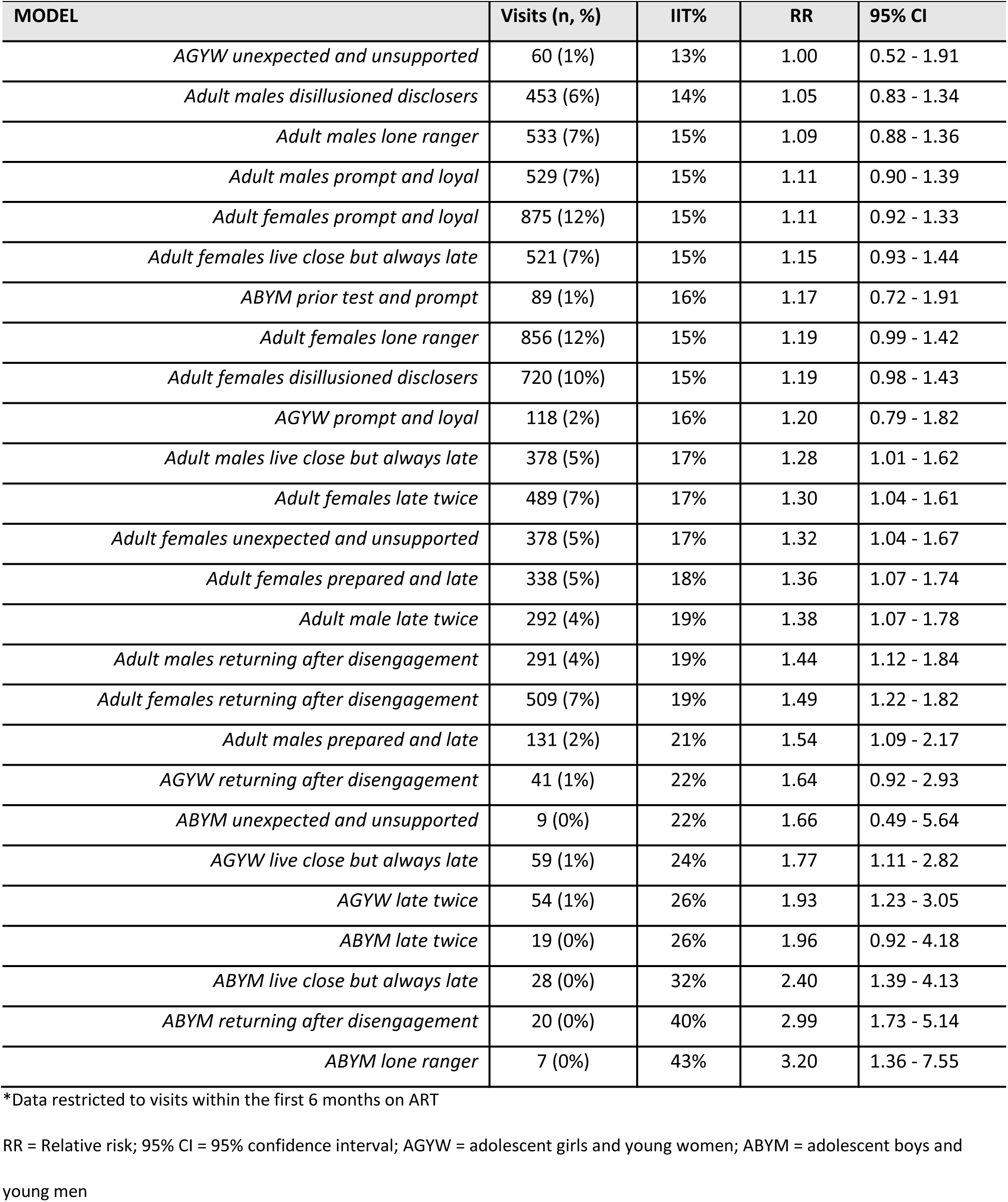
Proportion of visits classified as IIT stratified by archetype triaging approach and demographics (PREDICT and SLATE data)

| MODEL | Visits (n, %) | IIT% | RR | 95% CI |
| --- | --- | --- | --- | --- |
| **PREDICT DATA ARCHETYPES (N=925,639 VISITS)\*** |  |  |  |  |
| **All adult females (reference group)** | **572,154 (57%)** | **10%** | **Reference** |  |
| *Adult females prompt and loyal* | 384 316 (39%) | 8% | 0.83 | 0.82-0.84 |
| *Adult males prompt and loyal* | 173 750 (17%) | 9% | 0.91 | 0.89-0.92 |
| *ABYM prompt and loyal* | 6 511 (1%) | 10% | 0.96 | 0.89-1.03 |
| *AGYW prompt and loyal* | 43 938 (4%) | 13% | 1.29 | 1.26-1.32 |
| *Adult females late twice* | 51 244 (5%) | 15% | 1.47 | 1.44 - 1.51 |
| *Adult females shopper no-number* | 40 627 (4%) | 16% | 1.56 | 1.53 - 1.60 |
| *Adult males late twice* | 30 554 (3%) | 17% | 1.65 | 1.60 - 1.69 |
| *Adult males shopper no-number* | 18 945 (2%) | 17% | 1.70 | 1.65 - 1.76 |
| *ABYM shopper no-number* | 667 (0%) | 18% | 1.78 | 1.52 - 2.09 |
| *ABYM late twice* | 1 190 (0%) | 18% | 1.79 | 1.59 - 2.02 |
| *AGYW late twice* | 7 371 (1%) | 21% | 2.07 | 1.98 - 2.16 |
| *AGYW shopper no-number* | 4 194 (0%) | 22% | 2.11 | 1.99 - 2.24 |
| *Adult females returning after disengagement* | 19 784 (2%) | 23% | 2.29 | 2.23 - 2.35 |
| *Adult males returning after disengagement* | 9 874 (1%) | 26% | 2.56 | 2.47 - 2.65 |
| *AGYW returning after disengagement* | 3 639 (0%) | 30% | 2.90 | 2.75 - 3.05 |
| *ABYM returning after disengagement* | 426 (0%) | 31% | 3.07 | 2.66 - 3.53 |
| **SLATE DATA ARCHETYPES (N=7,199 VISITS)** |  |  |  |  |
| **All adult females (reference group)** | **4,141 (57%)** | **13%** | **Reference** |  |
| *AGYW prior test and prompt* | 52 (1%) | 6% | 0.43 | 0.14 - 1.30 |
| *AGYW super green* | 49 (1%) | 6% | 0.46 | 0.15 - 1.37 |
| *ABYM prompt and loyal* | 30 (0,5%) | 7% | 0.50 | 0.13 - 1.90 |
| *Adult females super green* | 1 418 (20%) | 10% | 0.67 | 0.56 - 0.80 |
| *Adult males super green* | 698 (10%) | 11% | 0.79 | 0.63 - 0.99 |
| *AGYW lone ranger* | 82 (1%) | 11% | 0.82 | 0.44 - 1.52 |
| *ABYM super green* | 148 (2%) | 12% | 0.91 | 0.59 - 1.41 |
| *Adult males unexpected and unsupported* | 370 (5%) | 12% | 0.91 | 0.68 - 1.21 |
| *Adult males prior test and prompt* | 636 (9%) | 12% | 0.92 | 0.73 - 1.14 |
| *Adult females prior test and prompt* | 1 012 (14%) | 13% | 0.97 | 0.81 - 1.17 |
| *AGYW unexpected and unsupported* | 60 (1%) | 13% | 1.00 | 0.52 - 1.91 |
| *Adult males disillusioned disclosers* | 453 (6%) | 14% | 1.05 | 0.83 - 1.34 |
| *Adult males lone ranger* | 533 (7%) | 15% | 1.09 | 0.88 - 1.36 |
| *Adult males prompt and loyal* | 529 (7%) | 15% | 1.11 | 0.90 - 1.39 |
| *Adult females prompt and loyal* | 875 (12%) | 15% | 1.11 | 0.92 - 1.33 |
| *Adult females live close but always late* | 521 (7%) | 15% | 1.15 | 0.93 - 1.44 |
| *ABYM prior test and prompt* | 89 (1%) | 16% | 1.17 | 0.72 - 1.91 |
| *Adult females lone ranger* | 856 (12%) | 15% | 1.19 | 0.99 - 1.42 |
| *Adult females disillusioned disclosers* | 720 (10%) | 15% | 1.19 | 0.98 - 1.43 |
| *AGYW prompt and loyal* | 118 (2%) | 16% | 1.20 | 0.79 - 1.82 |
| *Adult males live close but always late* | 378 (5%) | 17% | 1.28 | 1.01 - 1.62 |
| *Adult females late twice* | 489 (7%) | 17% | 1.30 | 1.04 - 1.61 |
| *Adult females unexpected and unsupported* | 378 (5%) | 17% | 1.32 | 1.04 - 1.67 |
| *Adult females prepared and late* | 338 (5%) | 18% | 1.36 | 1.07 - 1.74 |
| *Adult male late twice* | 292 (4%) | 19% | 1.38 | 1.07 - 1.78 |
| *Adult males returning after disengagement* | 291 (4%) | 19% | 1.44 | 1.12 - 1.84 |
| *Adult females returning after disengagement* | 509 (7%) | 19% | 1.49 | 1.22 - 1.82 |
| *Adult males prepared and late* | 131 (2%) | 21% | 1.54 | 1.09 - 2.17 |
| *AGYW returning after disengagement* | 41 (1%) | 22% | 1.64 | 0.92 - 2.93 |
| *ABYM unexpected and unsupported* | 9 (0%) | 22% | 1.66 | 0.49 - 5.64 |
| *AGYW live close but always late* | 59 (1%) | 24% | 1.77 | 1.11 - 2.82 |
| *AGYW late twice* | 54 (1%) | 26% | 1.93 | 1.23 - 3.05 |
| *ABYM late twice* | 19 (0%) | 26% | 1.96 | 0.92 - 4.18 |
| *ABYM live close but always late* | 28 (0%) | 32% | 2.40 | 1.39 - 4.13 |
| *ABYM returning after disengagement* | 20 (0%) | 40% | 2.99 | 1.73 - 5.14 |
| *ABYM lone ranger* | 7 (0%) | 43% | 3.20 | 1.36 - 7.55 |

\*Data restricted to visits within the first 6 months on ART
RR = Relative risk; 95% CI = 95% confidence interval; AGYW = adolescent girls and young women; ABYM = adolescent boys and young men

The development of the behavioural archetypes provided a more granular characterization of risk within each demographic stratum compared to a single risk estimate for any one demographic group. Figure 2 offers a visual depiction of how the point estimates for risk of treatment interruption vary when stratifying risk using demographic characteristics only compared to stratifying risk by combined demographic and behavioural characteristics. For both PREDICT and SLATE datasets, when risk of IIT is stratified by demographic characteristics only (gender and age), we find estimates of risk tend to cluster close together. In the SLATE datasets, for example, risk of IIT ranged from a relative risk of 0.97 (95% CI 0.63-1.49) for adolescent boys and young men to a relative risk of 1.15 (95% CI 0.91-1.46) among adolescent girls and young women; suggesting adolescent boys to be at similar risk for IIT compared to adult women. However, when the behavoural archetypes are considered within a singular demographic stratum (in this case, restricting to adolescent boys and young men), the point estimates for relative risk of treatment interruption at next scheduled visit spans a much wider range and subgroups with varying risk of IIT are revealed; characterized largely by prior visit attendance. The behavioural archetypes indicate that adolescent boys and young men who have attended clinic visits on time are at low risk of IIT at next visit compared to adult females, while those who have attended visits late at least twice in the past are twice as likely to experience treatment interruption (RR= 2.16; 95% CI 1.91-2.44; PREDICT datasets) and those who have previously disengaged from care are three times as likely to interrupt treatment at next scheduled visit (RR = 3.07; 95% CI 2.66 - 3.53; PREDICT datasets).

**Figure 2:**
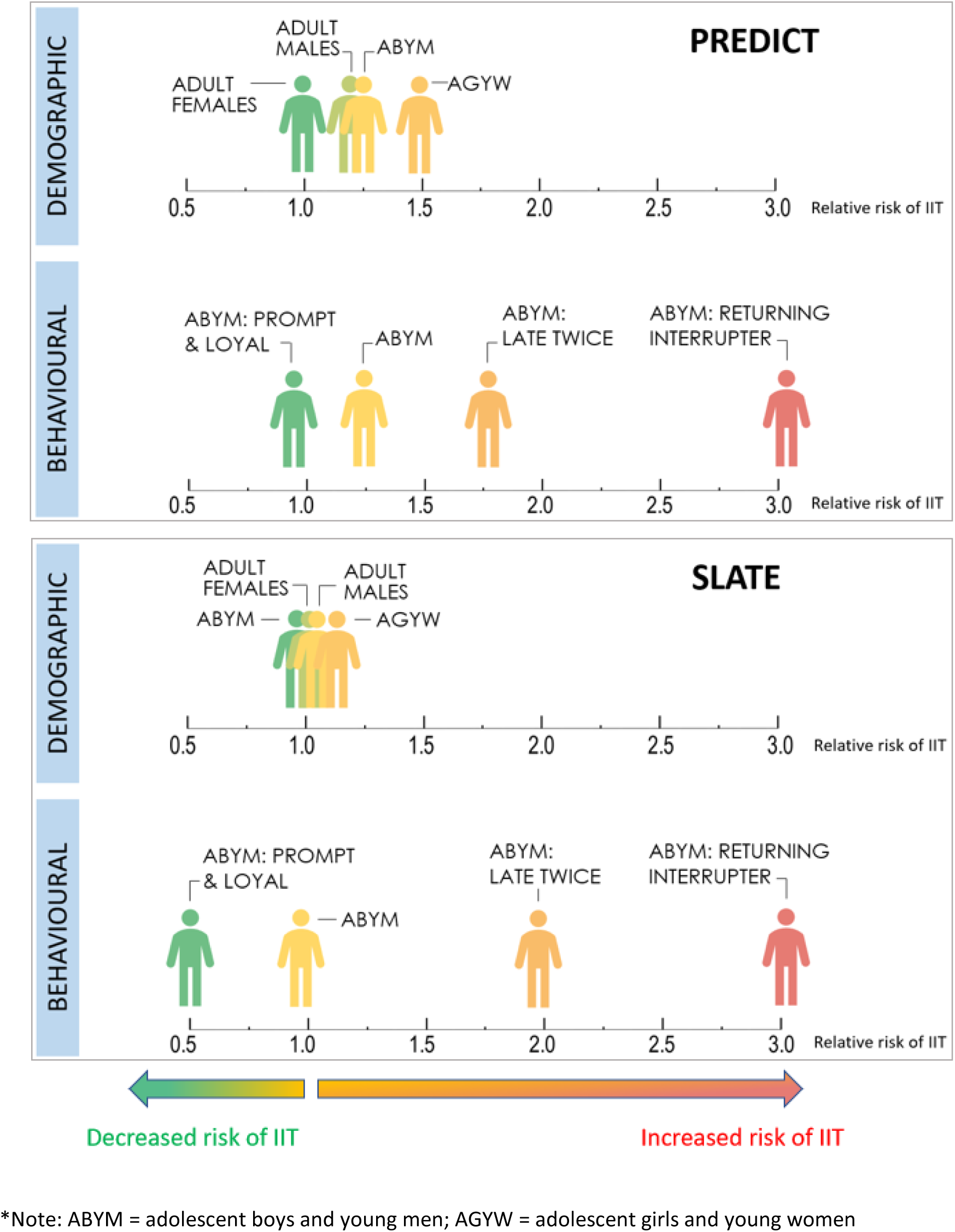
Relative risk of interruption in treatment (IIT) at next scheduled visit stratified by risk archetype and demographic strata.

## DISCUSSION

As HIV service delivery models expand and evolve, ensuring sustained client retention after treatment initiation remains a key priority. Many interventions to address disengagement from care are either applied universally to all clients engaged in treatment programs or reactively after clients have disengaged from care [28,29]; both scenarios utilize resources that do little to improve program outcomes. In this paper we present a novel application of machine learning and predictive algorithms to develop approaches that identify clients who are at heightened risk of disengagement before they experience interruptions in care or disengage entirely. Both of our models, if used in the course of a routine clinic visit, would allow healthcare providers to target interventions substantially more accurately than is currently possible, potentially improving retention among those at risk for disengagement if suitable interventions to address underlying drivers of risk are available as part of routine care.

Both of the approaches we report were successful, though in different ways. We note first that the original PREDICT model reproduced well in the SLATE datasets. Despite the differences in population, geography, and sample size, the results were very similar in terms of model performance metrics between the PREDICT and SLATE models. Across varying approaches and classification algorithms, the models were able to consistently predict approximately two in three visits classified as treatment interruptions.

The threshold approach is also useful for categorizing client groups at functional risk levels and offers an opportunity to triage clients for different intensity interventions. For example, our results confirm that rates of IIT are high for some clients during the first 6 months but are not universally so for all clients. The threshold approach to triaging was able to identify sub-groups of early ART clients most at risk for a treatment interruption (red group) and others at low risk (green group), an approach that can be readily interpreted and easily adapted into a simple scoring system for use at point-of-care. This could allow for triaging of clients at the facility level into high or low intensity models of service delivery before clients are eligible for existing differentiated service delivery models. Given that low risk visits comprised half of all visits and were associated with very low rates of IIT (6%), shifting clinician time and facility resources to higher risk clients could translate into important gains in efficiency without compromising quality of care for low risk groups.

The second approach, the archetype approach to risk triaging confirmed several important points. First, patterns of visit attendance are key in identifying risk of IIT in both directions, regardless of demographic sub-group. Of the three socio-behavioural risk groups with the lowest rates of IIT, two were characterized by on-time visit attendance (“super green” and “prior test and prompt”). In contrast, the three archetypes at highest risk of IIT (“returning after disengagement”, “live close but always late” and “prepared and late”) were all characterized by a history of late appointments or prior disengagement in care. This suggests that client behavior, as revealed by visit attendance, tends to be consistent and may present opportunities to intervene prior to disengagement among those who are at higher risk. It also allows providers to identify low risk groups who not only represent an important share of clients attending facility visits (32% of visits were among clients characterized as “super green”) but could potentially be safely managed with lower intensity models of care immediately after ART initiation, allowing for reallocation of time and resources to groups identified as priority risk groups. The use of behavioural archetypes allows for a more granular and detailed characterization of risk within a particular demographic profile (Figure 2). When risk of IIT is estimated for each of the behavoural archetypes within a singular demographic stratum, key sub-groups at increased risk of IIT are revealed; again, characterized largely by prior visit attendance. In this way, adolescent boys and young men simultaneously represent both the group at lowest risk of IIT at next visit (ABYM who have attended visits at the originating facility on time) as well as the group at nearly the highest risk of IIT (ABYM returning after previously disengaging from care). The behavioural archetype approach allows for identification of subgroups that have similar demographic characteristics but likely require quite different intervention strategies to support continuous engagement in care. As the information required to profile a client into a behavioural archetype is readily available at point-of-care, this approach may offer the potential to tailor interventions to specific groups in a more targeted way than has been available previously.

Where social and behavioural data are available, utilizing the archetype approach can also contribute to understanding not only particular client subgroups that are at risk of treatment interruption but also insight into the mechanisms underlying the increased risk. For example, when the socio-behavioural archetypes are stratified by demographic profiles (Table 5) we see a higher risk of IIT for AGYW who are classified as disillusioned disclosers and ABYM meeting the lone ranger archetype. Both of these archetypes are characterized by living alone, which suggests that young persons living with HIV may be vulnerable to a lack of social support as they navigate their HIV care journey. This knowledge could inform service delivery models providing differentiated care to this age group. In addition, we noted that while youth are generally at higher risk of treatment interruption, the subgroup of youth who reported being employed and had a visit scheduled within 7 days of payday were an archetype with one of the lowest risk of a subsequent treatment interruption (RR=1.13, 95% CI 0.81-1.57; Table 4), suggesting that scheduling of clinic visits may important for successful attendance among those with work commitments or where access to money for transport is key.

Finally, where the archetype approach to risk triaging provides insight into underlying drivers of risk of treatment interruption, it also creates the opportunity to map appropriate interventions to groups of ART clients most likely to benefit from them (Figure 3). For example, archetypes characterized by a lack of social support might be offered a treatment buddy or coach to assist them in establishing care during the early treatment period. Alternatively, a health worker might consider offering the choice of appointment scheduling to the employed youth – those who struggle to attend near payday because of work commitments might prefer a visit date earlier in the month, while another youth who needs their wages for transport money may prefer a visit scheduled shortly after payday. Used in this way, risk triaging offers an opportunity to optimize the impact of retention interventions by offering them to those among whom such interventions are most likely to have a positive impact on visit attendance while also reducing unnecessary resource expenditure by not offering the same interventions to clients who may neither want nor need them.

**Figure 3:**
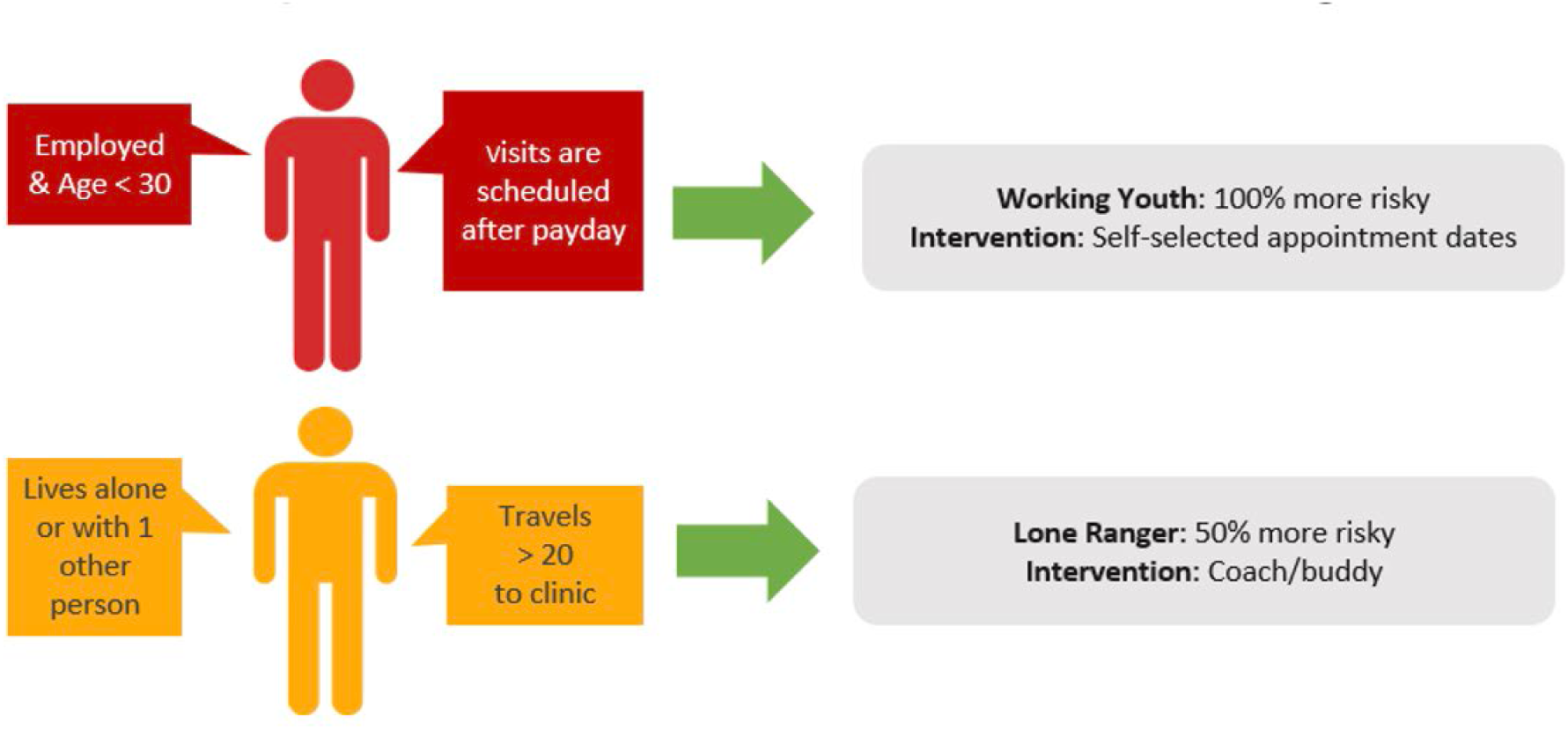
Schematic of intervention mapping guided by behavioural archetype.

Our results should be interpreted in light of the limitations in both the data sources used in this analysis and our approaches. Importantly, both data sources only observed clinic visits at the originating site. Clients may have attended HIV care at another facility but not been observed in the analytic datasets presented here; such visits may be misclassified as “not attended”. This is a common drawback to studies that rely on unlinked clinic records, as we did in both our analytic approaches (threshold and archetype) [26]. To the extent that this happened, our results will overestimate proportions of clinic visits not attended. The archetype results reflect several other limitations as well. Development of the client archetype approach is limited to the variables that were collected in the SLATE trials; other important archetypes may exist that we are unable to describe due to this limitation. The archetyping approach itself may be limited by the need for variables that are not currently routinely collected, though information such as whether a client lives alone or not could easily be added to routine data collection forms if it proved of sufficient value. Finally, generalizability of the archetype approach in particular may be limited to South Africa, as it relies on social and behavioral variables that vary by geographic location or culture.

## CONCLUSIONS

Despite the limitations mentioned above, our results have several important implications for HIV service delivery. First, the threshold risk triaging approach allows for identifying clients at risk of treatment interruptions while the archetype approach could potentially inform underlying obstacles to visit adherence and possible intervention mapping efforts. Of note is that these processes are intended to happen at point of care among clients still engaged in HIV health services, avoiding resource-intense tracing activities. We also present results and risk profiling relevant to the first six months after ART initiation; a key period for disengagement in care where the optimal model of service delivery remains unclear. Future work should address changes in risk states identified through these triaging approaches and the implications for long-term retention on ART. Finally, beyond improving the accuracy of risk prediction, our results represent an important step in introducing the results of machine learning and predictive analytic risk profiling into a routine practice setting. A simple tool for healthcare providers to utilize at point of care, before clients experiences negative outcomes, may be feasible using the characteristics found to be most predictive of future ITT in this study.

## Data Availability

All data results produced in the present study are contained in the manuscript and supplementary material. Source data for the SLATE model are available online at Boston University's data repository. Source data for the PREDICT models are owned by the South African Government and were used under license for the current study. Access to these is subject to restrictions owing to privacy and ethics policies set by the South African Government, so they are not publicly available. Requests to access these should be directed
to.

https://open.bu.edu/handle/2144/44321

## SUPPLEMENTARY TABLES AND FIGURES

Supplementary Table 1: Model building and validation process

Supplementary Table 2: Proportion visits with IIT at next scheduled visit stratified by current visit risk triaging classification and time on ART (varying threshold classification and model approaches)

## AUTHOR CONTRIBUTIONS

Conceptualization: MM, KSS, LDV

Methodology: MM, KSS, LDV, SP

Validation: KSS, LDV, SP

Software: SP, KSS, LDV

Formal analysis: MM, KSS, LDV, SP

Investigation: MM, KSS, LDV, SP

Data curation: PP, TC, AG, SR

Supervision: MM and SR

Original draft preparation: MM and SP

Review and editing: All authors

## REFERENCES

1. Keene CM, Ragunathan A, Euvrard J, English M, McKnight J, Orrell C. Measuring patient engagement with HIV care in sub-Saharan Africa: a scoping study. Journal of the International AIDS Society. 2022;25: 1–15. doi:10.1002/jia2.26025

2. Ford N, Geng E, Ellman T, Orrell C, Ehrenkranz P, Sikazwe I, et al. Emerging priorities for HIV service delivery. PLoS Medicine. 2020;17: 1–13. doi:10.1371/JOURNAL.PMED.1003028

3. NCT04429061. Reaching 90 90 90 in Adolescents in Zambia: using All Our SKILLZ. https://clinicaltrials.gov/show/NCT04429061. 2020.

4. Beres LK, Denison JA, Schwartz S, Simbeza S, Mwamba C, Sikombe K, et al. Patterns and Predictors of Incident Return to HIV Care Among Traced, Disengaged Patients in Zambia: Analysis of a Prospective Cohort. Journal of acquired immune deficiency syndromes (1999). 2021;86: 313–322. doi:10.1097/QAI.0000000000002554

5. Tweya H, Gareta D, Chagwera F, Ben-Smith A, Mwenyemasi J, Chiputula F, et al. Early active follow-up of patients on antiretroviral therapy (ART) who are lost to follow-up: the “Back-to-Care” project in Lilongwe, Malawi. Tropical medicine & international health : TM & IH. 2010;15 Suppl 1: 82–89. doi:10.1111/j.1365-3156.2010.02509.x

6. Satti H, McLaughlin MM, Omotayo DB, Keshavjee S, Becerra MC, Mukherjee JS, et al. Outcomes of Comprehensive Care for Children Empirically Treated for Multidrug-Resistant Tuberculosis in a Setting of High HIV Prevalence. Madhi SA, editor. PLoS ONE. 2012;7: e37114. doi:10.1371/journal.pone.0037114

7. Bershetyn A, Odeny TA, Lyamuya R, Nakiwogga-Muwanga A, Diero L, Bwana M, et al. The causal effect of tracing by peer health workers on return to clinic among patients who were lost to follow-up from antiretroviral therapy in Eastern Africa: A “natural experiment” arising from surveillance of lost patients. Clinical Infectious Diseases. 2017;64: 1547–1554. doi:10.1093/cid/cix191

8. Etoori D, Wringe A, Renju J, Kabudula CW, Gomez-Olive FX, Reniers G. Challenges with tracing patients on antiretroviral therapy who are late for clinic appointments in rural South Africa and recommendations for future practice. Global Health Action. 2020;13. doi:10.1080/16549716.2020.1755115

9. USAID. Data and Advanced Analytics in HIV Service Delivery: Use Cases to Help Reach 95-95-95. 2020.

10. Kerschberger B, Aung A, Mpala Q, Ntshalintshali N, Mamba C, Schomaker M, et al. Predicting, Diagnosing, and Treating Acute and Early HIV Infection in a Public Sector Facility in Eswatini. J Acquir Immune Defic Syndr. 2021;88: 506–517.

11. Giovenco D, Pettifor A, MacPhail C, Kahn K, Wagner R, Piwowar-Manning E, et al. Assessing risk for HIV infection among adolescent girls in South Africa: an evaluation of the VOICE risk score (HPTN 068). J Int AIDS Soc. 2019;22: e25359.

12. Brown LB, Miller WC, Kamanga G, Kaufman JS, Pettifor A, Dominik RC, et al. Predicting partner HIV testing and counseling following a partner notification intervention. AIDS Behav. 2012;16: 1148–55. doi:10.1007/s10461-011-0094-9

13. Stevens WS, Gous NM, Macleod WB, Long LC, Variava E, Martinson NA, et al. Multidisciplinary point-of-care testing in south african primary health care clinics accelerates HIV ART initiation but does not alter retention in care. J Acquir Immune Defic Syndr (1988). 2017;76: 65–73. doi:10.1097/QAI.0000000000001456

14. Auld AF, Fielding K, Agizew T, Maida A, Mathoma A, Boyd R, et al. Risk scores for predicting early antiretroviral therapy mortality in sub-Saharan Africa to inform who needs intensification of care: a derivation and external validation cohort study. BMC Med. 2020;18: 1–19.

15. Maskew M, Brennan AT, Fox MP, Vezi L, Venter WDF, Ehrenkranz P, et al. A clinical algorithm for same-day HIV treatment initiation in settings with high TB symptom prevalence in South Africa: The SLATE II individually randomized clinical trial. PLoS Med. 2020;17. doi:10.1371/JOURNAL.PMED.1003226

16. Mamo DN, Yilma TM, Fekadie M, Sebastian Y, Bizuayehu T, Melaku MS, et al. Machine learning to predict virological failure among HIV patients on antiretroviral therapy in the University of Gondar Comprehensive and Specialized Hospital, in Amhara Region, Ethiopia, 2022. BMC Med Inform Decis Mak. 2023;23. doi:10.1186/s12911-023-02167-7

17. Frijters EM, Hermans LE, Wensing AMJ, Devillé WLJM, Tempelman HA, De Wit JBF. Risk factors for loss to follow-up from antiretroviral therapy programmes in low-income and middle-income countries. AIDS. 2020;34: 1261–1288. doi:10.1097/QAD.0000000000002523

18. Makurumidze R, Decroo T, Jacobs BKM, Rusakaniko S, Van Damme W, Lynen L, et al. Attrition one year after starting antiretroviral therapy before and after the programmatic implementation of HIV “Treat All” in Sub-Saharan Africa: a systematic review and meta-analysis. BMC Infectious Diseases. 2023;23: 1–13. doi:10.1186/s12879-023-08551-y

19. Conan N, Simons E, Chihana ML, Ohler L, FordKamara E, Mbatha M, et al. Increase in HIV viral suppression in KwaZulu-Natal, South Africa: Community-based cross sectional surveys 2018 and 2013. What remains to be done? PLoS One. 2022;17. doi:10.1371/journal.pone.0265488

20. Maskew M, Sharpey-Schafer K, De Voux L, Crompton T, Bor J, Rennick M, et al. Applying machine learning and predictive modeling to retention and viral suppression in South African HIV treatment cohorts. Scientific Reports. 2022;12: 12715. doi:10.1038/s41598-022-16062-0

21. UNAIDS Joint United Nations Programme on HIV/AIDS. Global AIDS Update 2021: Confronting Inequalities. Geneva, Switzerland; 2021.

22. Rosen S, Maskew M, Larson BA, Brennan AT, Tsikhutsu I, Fox MP, et al. Simplified clinical algorithm for identifying patients eligible for same-day HIV treatment initiation (SLATE): Results from an individually randomized trial in South Africa and Kenya. Newell M-L, editor. PLOS Medicine. 2019;16: e1002912. doi:10.1371/journal.pmed.1002912

23. Maskew M, Sharpey-Schafer K, De Voux L, Crompton T, Bor J, Rennick M, et al. Applying machine learning and predictive modeling to retention and viral suppression in South African HIV treatment cohorts. Scientific Reports. 2022;12: 12715. doi:10.1038/s41598-022-16062-0

24. Esra R, Carstens J, Le Roux S, Mabuto T, Eisenstein M, Keiser O, et al. Validation and improvement of a machine learning model to predict interruptions in antiretroviral treatment in South Africa. Journal of acquired immune deficiency syndromes (1999). 2022. doi:10.1097/QAI.0000000000003108

25. Maskew M, Brennan AT, Venter WDF, Fox MP, Vezi L, Rosen S. Retention in care and viral suppression after same-day ART initiation: One-year outcomes of the SLATE I and II individually randomized clinical trials in South Africa. Journal of the International AIDS Society. 2021;24: e25825. doi:10.1002/jia2.25825

26. Maskew M, Benade M, Huber A, Pascoe S, Sande L, Malala L, et al. Patterns of engagement in care during clients’ first 12 months after HIV treatment initiation in South Africa: A retrospective cohort analysis using routinely collected data. PLOS Global Public Health. 2024;4: e0002956. doi:10.1371/journal.pgph.0002956

27. Pedregosa F, Varoquaux G, Gramfort A, Michel V, Prettenhofer P, Weiss R, et al. Scikit-learn: Machine Learning in Python. Journal of Machine Learning Research. 2011;12: 2825–2830.

28. Long L, Kuchukhidze S, Pascoe S, Nichols BE, Fox MP, Cele R, et al. Retention in care and viral suppression in differentiated service delivery models for HIV treatment delivery in sub-Saharan Africa: a rapid systematic review. Journal of the International AIDS Society. John Wiley and Sons Inc; 2020. doi:10.1002/jia2.25640

29. Rosen S, Grimsrud A, Ehrenkranz P, Katz I. Models of service delivery for optimizing a patient’s first six months on antiretroviral therapy for HIV: An applied research agenda. Gates Open Research. 2020;4: 1–15. doi:10.12688/gatesopenres.13159.1

