## Supplementary tables and figures for "Triaging clients at risk of disengagement from HIV care: Application of a predictive model to clinical trial data in South Africa"

### Supplementary Table 1: Model building and validation process

The PREDICT and SLATE model output presented in this manuscript used the AdaBoost (adaptive boosting binary classification) algorithm from scikit-learn. This boosting method was selected as it previously demonstrated better performance metrics than logistic regression during a validation study and the slight improvement from using gradient boosting methods did not justify the complexity added to interpretation by this approach. These prior validation efforts by Esra et al were conducted using datasets extracted from the same electronic patient records as the PREDICT model, Tier.net from the Gauteng and North West provinces of South Africa (GA/NW datasets). The datasets used by Esra et al. were collected as part of routine patient care and represented large sample sizes (>900,000 and >300,000 patients for the PREDICT and GA/NW datasets, respectively). Despite almost identical performance of the model in both datasets (sensitivity of 60% and 61% for the PREDICT and GA/NW datasets, respectively), it remained unclear how model performance would be affected by a reduced sample size and data collected outside of routine settings.

As with the PREDICT model building process, our approach commenced by separating the SLATE dataset into training and test sets. Training sets are datasets with known exposure and outcome variables used in machine learning approaches to allow the algorithm to "learn" the predictive importance of exposure variables in terms of correctly classifying each specified outcome. For test sets, the exposure variables are separated from the outcome variables (unseen) and given to the final classifier algorithm. The model is tested on this unseen data set by generating predicted outcomes using the predictor variables from the unseen test set. These predicted outcomes are then compared to the known outcomes in the test set and the model is scored according to standard performance metrics. Supplementary Table 1 describes each model's performance metrics in terms of sensitivity, specificity, positive and negative predictive value, accuracy and area under the curve. We also compare these performance metrics of the SLATE model to the original PREDICT model overall and the model restricted to the first 6 months on ART (the treatment period relevant during the SLATE trials) as well as results from using a gradient boosting approach to the SLATE data.

| | **PREDICT model** | **SLATE model (all variables plus up-sampled visit data)** | ****SLATE model (all variables)** | **SLATE model (variables available in PREDICT model+ upsampled visit data)** | ****SLATE model (limited to variables available in PREDICT model)** |
|---|---|---|---|---|---|
| Sample size (test set) | N = 1,399,145 patient visits | n = 13,458 patient visits | n = 11,400 patient visits | n = 8,487 patient visits | n = 7,199 patient visits |
| Algorithm | AdaBoost | GradientBoosting | GradientBoosting | GradientBoosting | GradientBoosting |
| Accuracy | 66% (n = 1,399,145) | 69% (n = 1,698) | 66% (n = 1,440) | 63% (n = 1,698) | 65% (n = 1,440) |

| | | | | | |
|---|---|---|---|---|---|
| Sensitivity | 61% (n = 146,881) | 58% (n = 245) | 52% (n = 200) | 51% (n = 245) | 52% (n = 200) |
| Specificity | 67% (n = 1,252,264) | 71% (n = 1,453) | 69% (n = 1,240) | 65% (n = 1,453) | 68% (n = 1,240) |
| Positive predictive value | 18% (n = 503,730 total positive predictions) | 25% (n = 563 total positive predictions) | 21% (n = 490 total positive predictions) | 19% (n = 636 total positive predictions) | 21% (n = 506 total positive predictions) |
| Negative predictive value | 94% (n = 895,415 total negative predictions) | 91% (n = 1,135 total negative predictions) | 90% (n = 934 total negative predictions) | 89% (n = 1,062 total negative predictions) | 90% (n = 950 total negative predictions) |
| AUC | 0.688 | 0.676 | 0.6512 | 0.601 | 0.6224 |
| Algorithm | - | AdaBoost | AdaBoost | AdaBoost | AdaBoost |
| Accuracy | - | 65% (n = 1,698) | 63% (n = 1,440) | 61% (n = 1,698) | 61% (n = 1,440) |
| Sensitivity | - | 57% (n = 216) | 52% (n = 200) | 50% (n = 245) | 55% (n = 200) |
| Specificity | - | 67% (n = 1,453) | 64% (n = 1,240) | 63% (n = 1,453) | 61% (n = 1,240) |
| Positive predictive value | - | 22% (n = 626 total positive predictions) | 19% (n = 544 total positive predictions) | 18% (n = 666 total positive predictions) | 19% (n = 589 total positive predictions) |
| Negative predictive value | - | 90% (n = 1,072 total negative predictions) | 89% (n = 896 total negative predictions) | 88% (n = 1,032 total negative predictions) | 89% (n = 851 total negative predictions) |
| AUC | - | 0.663 | 0.614 | 0.609 | 0.603 |

**Supplementary Table 2: Proportion visits with IIT at next scheduled visit stratified by current visit risk triaging classification and time on ART (varying threshold classification and model approaches)**

| **SOURCE** | **PREDICT DATASETS** | | | | | **SLATE DATASETS** | | |
|---|---|---|---|---|---|---|---|---|
| **TIME ON ART** | **OVERALL** | **FIRST VISIT** | | **0-6 MONTHS ON ART** | | **OVERALL** | **FIRST VISIT** | **0-6 MONTHS ON ART** |
| **Algorithm** | AdaBoost | Gradient Boosted | AdaBoost | Gradient Boosted | AdaBoost | AdaBoost | AdaBoost | AdaBoost |
| **Total N** | 1,399,145 | 41,751 | 10,205 | 194,469 | 63,377 | 1,440 | 155 | 791 |
| **Test set N** | 146,881 (11%) | 7,979 (19%) | 5,043 (49%) | 19,583 (10%) | 16,873 (27%) | n=200 (14%) | n=34 (22%) | n=114 (14%) |
| **Model predicted:**<br>**IIT**<br>**No IIT** | **Observed data:**<br>**IIT No IIT**<br>837674 57741<br>414590 89140 | **Observed data:**<br>**IIT No IIT**<br>21729 3652<br>12043 4327 | **Observed data:**<br>**IIT No IIT**<br>3845 3267<br>1317 1776 | **Observed data:**<br>**IIT No IIT**<br>113191 7766<br>61695 11817 | **Observed data:**<br>**IIT No IIT**<br>30361 5473<br>16143 11400 | **Observed data:**<br>**IIT No IIT**<br>799 97<br>441 103 | **Observed data:**<br>**IIT No IIT**<br>81 40<br>21 13 | **Observed data:**<br>**IIT No IIT**<br>439 238<br>60 54 |
| **Accuracy** | 66% | 62% | 55% | 64% | 66% | 63% | 61% | 62% |
| **Specificity** | 67% | 64% | 74% | 65% | 65% | 64% | 67% | 65% |
| **Sensitivity** | 61% | 54% | 35% | 60% | 68% | 52% | 38% | 47% |
| **Precision** | 18% | 26% | 57% | 16% | 41% | 19% | 25% | 19% |
| **AUC** | 0.688 | 0.633 | 0.563 | 0.674 | 0.713 | 0.614 | 0.575 | 0.572 |
